# Suriname Cerebral Palsy Register – preliminary data from the first hospital-based surveillance of children with cerebral palsy in Suriname

**DOI:** 10.1101/2023.09.01.23294913

**Authors:** Marlies Declerck, Israt Jahan, Neirude PA Lissone, Fenna Walhain, Ruby Chin A Fat, Marianne Fleurkens, Sarfaraz H. J. Muradin, Rhea Cruden, Els Ortibus, Nadia Badawi, Gulam Khandaker

**Author notes:** **Corresponding author:** Marlies Declerck, Faculteit der Medische Wetenschappen, Kernkampweg 5,00000 Paramaribo, Suriname.

## Abstract

**Aim:** To describe the epidemiology of cerebral palsy (CP) among children and their access to rehabilitation and education in Suriname.

**Methods:** Hospital-based surveillance of children with CP at the Academic Hospital in Suriname between August 2018 and March 2020.

**Results:** Data were collected from 82 children with CP (mean (SD) age: 5y 0mo (3y 10mo)). The median age of diagnosis was 5y 2mo. Spastic CP was predominant (73.2%), 58.8% had gross motor function classification system level III-V. Overall, 42.2% had preterm birth compared to 13.9% reported nationally (p<0.001), 57% had birth-related complications in comparison to 15% reported nationally (p<0.001). Additionally, 39.1% had birth asphyxia, 23.2% had early feeding difficulties. Sixty-two percent were admitted to Neonatal Intensive Care Unit, 54.0% of them required ventilation. Majority (82.5%) had CP acquired pre-or perinatally and 17.5% had CP acquired post-neonatally. Sixteen percent had never received any rehabilitation services, and 31.9% of the school-aged children were not enrolled in any education system.

**Interpretation:** The high burden of known risk factors, substantially delayed diagnosis and severe functional impairment among registered children with CP in Suriname is concerning. Public health interventions targeting early diagnosis and intervention could improve the functional outcome of children with CP in Suriname.

**What this paper adds:**

⍰ The epidemiology of cerebral palsy (CP) among children in Suriname using data from the first hospital-based surveillance of CP in the country and region.
⍰ Children with CP in Surname are diagnosed too late for early intervention.
⍰ Most of the children with CP have potentially-preventable risk factors.
⍰ Majority of them have severe functional limitations.
⍰ Targeted actions for early diagnosis and intervention, rehabilitation and inclusive education for children with CP is required in Suriname.

## Introduction

Cerebral Palsy (CP), is”a group of permanent disorders of the development of movement and posture, causing activity limitations that are attributed to non-progressive disturbances that occurred in the developing foetal or infant brain”. (1) CP is one of the leading causes of childhood disability globally with an estimated prevalence of 1.6 per 1000 live births in high income countries (HICs) and a markedly higher prevalence in low- and middle-income countries (LMICs; estimated 2.9‒3.4 per 1000 children). (2, 3)

Data from LMICs show that most children with CP in LMICs have severe motor impairments, malnutrition, and low quality of life. (4-7) Furthermore, the common risk factors, timing of brain injury and clinical presentation differ between HICs and LMICs as well as across LMICs (4-7). For instance, the rate of preterm birth and low birthweight was reported to be 43% each among children with CP registered in the Australian CP Register. (8) CP registers in Bangladesh, Indonesia and Nepal reported that only 11-18% of the children with CP had preterm birth and 27-38% of children with CP had low birthweight. (4, 9, 10) Even within LMICs, the risk factors of CP vary substantially between different countries. Jahan et al., (6) found that the incidence of birth asphyxia was 65.4%, 38.3%, 36.5% and 26.9% among children with CP registered in Bangladesh, Nepal, Ghana and Indonesia, respectively. The vast differences between LMICs indicate the need for country-specific data on the epidemiology of CP in LMICs.

The Republic of Suriname is a middle-income country located on the northeast coast of South America. Approximately 70% of the total population (e.g., almost 570,000) is based in the urban-coastal area of Paramaribo, the capital of Suriname, and Wanica. (11) Suriname’s population is among the most varied in the world, including the Indigenous Amerindians, the Creoles and Maroons, the Hindustanis and Javanese, descendants from China, from a number of European and Middle Eastern countries, and more recently immigrants from various Latin American and Caribbean countries. (12)

The healthcare system in Suriname consists of a mixed system including three major types of financing i.e., the state health insurance fund (comprehensive package of health benefits for approximately 35% of the population, free of charge for civil servants and all children), the Ministry of Social Affairs (MSA) (free of charge for the poor and near poor, covering approximately 42% of the population), and private firms/health insurance (approximately 20% of the population). (13) The Surinamese people receive medical services from an array of public and private health care providers. The public providers of primary health care are the Regional Health Services (RGD), a state foundation, and the Medical Mission (MM), an NGO. Both institutions are subsidized by the Government. The RGD provides services to the poor and near-poor in the coastal area through 50 clinics (generally lower socioeconomic means), whereas the MM provides curative and preventive health services to the interior Maroon and Amerindian population through 50 health centers and health posts. (14) Furthermore, consultation bureaus of the public health care system are responsible for following up of the development of children until the age of one year, with one follow-up at the age of four years old. During these follow-ups, if a nurse detects a developmental delay or risk, they refer the child to the in-house General Practitioner (GP) and to the tertiary hospitals in the capital city Paramaribo for further assessments. If developmental issues are detected by the GP, they are referred to the rehabilitation centres in Paramaribo (e.g., Academic Hospital in Paramaribo) for follow-up and services. Except for the Maroon and Amerindian minorities in the interior, for families to access care from medical specialists, physical therapists (PT), occupation therapists (OT), speech therapy (ST), a referral from the general practitioners (GP) are essential. (14)

The public Academic Hospital (which is also a training institution), is one of the six hospitals that provides a variety of specialty services (both inpatient and ambulatory) in Suriname. The hospital employs the majority of medical specialists and offers nearly all types of specialty care. The Rehabilitation Center, as a subdivision of the Academic Hospital in Paramaribo, provides services to referral patients for the construction of artificial limbs, and physical, occupational, and speech therapy. With a staff of 14 paramedics and medical professionals, the Center registers approximately 6,000 patient visits each year. In 2004, the Center opened a special children’s unit under the care of a specialized physiotherapist. Since then, the centre has been serving as one of major sites for referral and specialized services for children with different forms of disabilities. This unit currently provides therapies and services to all children of the school for children with disabilities (Mytylschool) (mostly CP and muscular dystrophy). (14)

However, until 2018, there was no register for documentation and follow-up of children with CP and other neurodevelopmental disorders at the unit or in the country. According to the latest multiple indicator cluster survey (MICS-2018), 13% of children aged 5-17 years in Suriname have at least one type of functional difficulty (15). Whereas the disability rate was reported 23.7% among children aged 2-9 years in the earlier MICS study in Suriname.(16) Evidence from other LMICs and globally, indicates that CP is one leading causes of childhood disabilities especially those with mobility impairments. However, such information on CP (e.g., disease burden, aetiology and rehabilitation needs) is lacking in Suriname which hinders early diagnosis and adequate management of children with CP in the country.

CP register plays a crucial role in defining the epidemiology of CP as well as improving service provision, access to early diagnosis and early intervention through active follow-up and guidance. (17-20) Although, HICs like Europe and Australia established the largest CP registers more than 30 years ago, (3, 17, 18); LMICs such as Bangladesh, Nepal, Sri Lanka, Indonesia and Ghana have recently established a country-specific CP register. (4, 6, 7, 21) However, none existed in Suriname, nor in the Caribbean or South America until 2018, when our group established the first CP register in Suriname.

In this study, we aimed to report our preliminary findings from the Suriname CP Register (SURCPR) and describe the epidemiology of CP among children attending Academic Hospital in Paramaribo, Suriname.

## Methods

The SUR-CPR was established in 2018 by a group of leading researchers and service providers of children with CP in Suriname with support from international experts (e.g., the Australian Cerebral Palsy Register (ACPR) and the Bangladesh Cerebral Palsy Register (BCPR) groups). A hospital-based surveillance methodology was used for participant recruitment following a standard protocol.

### Study settings and participants

A hospital-based surveillance program for children with CP was set up at the Academic Hospital Paramaribo (Academisch Ziekenhuis Paramaribo or AZP). AZP is the largest of the six hospitals in Suriname and is located in the Paramaribo district. The district encompasses ∼182 square kilometres with ∼236,065 population (∼64,743 children aged <18y). (11, 22) 92.2% of all births in Suriname take place in hospitals/ healthcare facilities and an estimated 30% of those births take place at the AZP. (23, 24) The neonatal unit of AZP is the only tertiary referral centre to provide health care services to critically ill new-borns in Suriname. (24) As described in the earlier sections, the Rehabilitation Center and the special children’s unit of Academic Hospital in Paramaribo is a key referral point for rehabilitation services (e.g., artificial limbs, PT, OT, ST) to children (mostly CP and muscular dystrophy) and adults in the country. Children with suspected CP aged <18 years attending the Paediatric, Neurology and Rehabilitation services of the AZP were eligible to participate. The case definition of CP as described by the Surveillance of CP in Europe (SCPE) and the Australian CP Register (ACPR) was used for the SUR-CPR. (3, 17)

### Data collection/Procedure

Detailed medical and neurodevelopmental assessments were performed by a paediatrician or neurologist trained by international experts following standard guidelines. (3, 17) Children aged <5 years at the time of assessment were followed-up again after completing five years of age to confirm diagnosis and records were updated accordingly. A standardized clinical registration form adapted from the BCPR and SCPE was used to gather information on sociodemographic characteristics, risk factors, clinical description, nutrition, rehabilitation, and education status. Sociodemographic characteristics were collected according to the national standards. (22) This included: age and sex of the child with CP, living area, parental characteristics (e.g., age, ethnicity and educational level), household access to water supply, electricity and toilet facility, number of household members and monthly family income in local currency (i.e. Surinamese dollar or SRD). Information about known risk factors was collected based on available medical records and clinical history provided by the primary caregivers (see supplementary study material 1). Clinical description of CP was documented based on detailed assessment, review of the medical records and interviewing the primary caregivers at the time of registration. Documentation was made on age at CP diagnosis, the timing of brain injury causing CP, predominant motor type of CP, topography, and functional classification systems. Predominant motor type and topography were documented following the SCPE guidelines. (17) The Gross Motor Function Classification System Expanded and Revised (GMFCS E&R), (25) the Manual Ability Classification System (MACS), (26) and the Communication Function Classification System (CFCS) (27) were used to document gross motor function, capacity to handle objects and communication performance, respectively. Additionally, the presence of any associated impairment was documented if indicated in the medical records. Furthermore, the Eating and Drinking Ability Classification System (EDACS) (28) and the presence of dysphagia among the participating children were also documented based on the caregiver’s response.The rehabilitation status of all children and their enrolment in any educational system (for children aged 4 years or above) were documented. For rehabilitation status, we documented if a child ever received any rehabilitation services, if yes then we documented the type of services received (e.g., physiotherapy (PT), occupational therapy (OT), speech and language therapy (SLT)). Additionally, for children who ever received rehabilitation services, we documented the frequency of receiving any of those therapies (e.g., PT, OT, SLT) in six months prior to the registration. Receipt of any assistive devices was also documented.

### Ethics approval

Ethical approval was granted by the Medical Ethical Committee of the Ministry of Health, Suriname (VG34-17A). Informed written consent was obtained from parents or caregivers of all the study participants.

### Data management and statistical analysis

Data management and analysis were performed using SPSS (version 23). The sociodemographic and clinical characteristics, function classifications, access to rehabilitation and education were described. The mean values with standard deviation (SD) were reported for the continuous variables (i.e. age at assessment, monthly family income, birth weight, age of CP diagnosis) considering the data distribution. The Apgar score at 5 min was considered low if scored 7 or less (24). Low birthweight was documented if a child weighed <2500 grams (27). The gestational age was categorized into four groups (i.e. extremely preterm, very preterm, moderate to late preterm, and term) following the WHO cut-off values. (30) Valid percentages were documented. The proportion of different socio-demographic characteristics and known risk factors amongst children with CP were compared with the national or regional average (i.e. in the general population) or findings from any large-scale relevant study. Chi-squared test, fishers exact test or binomial test were used as appropriate. A *p*-value <0.05 was considered for statistical significance.

## Results

Between August 2018 and March 2020, 82 children (37 girls, 45 boys) with clinically confirmed CP were registered in the SUR-CPR with a mean (SD) age of 5 years (3.8y) ranging between 1 and 17 years at the time of registration.

### Socio-demographic characteristics

The socio-demographic data and details on biological parents (and caregivers) of children with CP registered into the SUR-CPR are summarized in **Table 1**. Overall, 87.8% children were aged below 10 years. Among the mothers, 34.1% were Maroon, 23.2% were Hindustani and 6.1% were Indigenous. A similar representation of different ethnicity was observed among the fathers of participating children. Overall, 12.5% of mothers did not receive any formal schooling and 30.3% of mothers had completed higher studies. Both these percentages were significantly higher than the national averages (*p* < 0.001 for both). Similar numbers were observed for paternal educational level. Most of the fathers (94.6%,) and nearly half of the mothers (49.4%) were involved in some income-generating activities (IGA). The majority of the caregivers (42.3%) had a monthly family income ranging between SRD 1500-3000 (USD 150-300 at the time of registration) and 33.3% had a monthly family income > SRD 3000 (>USD 300 at the time of registration).

**Table 1.** Socio-demographic characteristics of the participating children.

| Socio-demographic characteristics | n (%) | Regional/ national distribution, % | p value |
| --- | --- | --- | --- |
| Age at assessment in year, n=82 |  |  |  |
| 0-4 | 40 (48.8) | 28.0<sup>1</sup> | <0.001<sup>3</sup> |
| 5-9 | 32 (39.0) | 28.6<sup>1</sup> |  |
| 10-14 | 7 (8.5) | 26.8<sup>1</sup> |  |
| 15- <18 | 3 (3.7) | 16.7<sup>1</sup> |  |
| Mean (SD), 95% CI | 5.8 (3.8),<br>5.0-6.7 | N/A | N/A |
| Gender, n=82 |  |  |  |
| Male | 45 (54.9) | 51.2<sup>1</sup> | 0.04<sup>4</sup> |
| Female | 37 (45.1) | 48.8<sup>1</sup> |  |
| Ethnicity of mother, n=82 |  |  |  |
| Maroon | 28 (34.1) | 16.0<sup>2</sup> | <0.001<sup>5</sup> |
| Hindustani | 19 (23.2) | 30.0<sup>2</sup> | 0.11<sup>5</sup> |
| Creole | 14 (17.1) | 21.0<sup>2</sup> | 0.23<sup>5</sup> |
| Javanese | 10 (12.2) | 13.0<sup>2</sup> | 0.49<sup>5</sup> |
| Indigenous | 5 (6.1) | 2.0<sup>2</sup> | 0.02<sup>5</sup> |
| Mixed Ethnicity | 5 (6.1) | 16.0<sup>2</sup> | 0.01<sup>5</sup> |
| Chinese | 1 (1.2) | 1.0<sup>3</sup> | 0.56<sup>5</sup> |
| Ethnicity of father, n=82 |  |  |  |
| Maroon | 29 (35.4) | 15.0<sup>2</sup> | <0.001<sup>5</sup> |
| Hindustani | 18 (22.0) | 31.0<sup>2</sup> | 0.05<sup>5</sup> |
| Mixed ethnicity | 11 (13.4) | 16.0<sup>2</sup> | 0.32<sup>5</sup> |
| Creole | 10 (12.2) | 20.0<sup>2</sup> | 0.05<sup>5</sup> |
| Javanese | 10 (12.2) | 13.0<sup>2</sup> | 0.49<sup>5</sup> |
| Indigenous | 2 (2.4) | 1.0<sup>2</sup> | 0.20<sup>5</sup> |
| Chinese | 2 (2.4) | 2.0<sup>2</sup> | 0.49<sup>5</sup> |
| Maternal educational level, n=79 |  |  |  |
| No formal schooling | 10 (12.6) | 3.8<sup>1</sup> | <0.001<sup>5</sup> |
| GLO (primary school) | 19 (24.0) | 13.5<sup>1</sup> | 0.01<sup>5</sup> |
| LBGO (Junior Secondary Vocational School) | 13 (16.4) | 68.7<sup>1</sup> | <0.001<sup>5</sup> |
| MULO (Junior Secondary General School) | 13 (16.4) |  |  |
| VMBO (Pre-Vocational Secondary Education) | 10 (12.6) | 13.9<sup>1</sup> | <0.001<sup>5</sup> |
| HAVO/VWO (Pre-University Education) | 6 (7.6) |  |  |
| HBO/University | 8 (10.1) |  |  |
| Paternal educational level, n=61 |  |  |  |
| No formal schooling | 8 (13.1) | 1.7<sup>1</sup> | <0.001<sup>5</sup> |
| GLO (primary school) | 16 (26.2) | 18.0<sup>1</sup> | 0.30<sup>5</sup> |
| LBGO (Junior Secondary Vocational School) | 5 (8.2) | 71.3<sup>1</sup> | <0.001<sup>5</sup> |
| MULO (Junior Secondary General School) | 12 (19.7) |  |  |
| VMBO (Pre-Vocational Secondary Education) | 11 (18.0) | 8.3<sup>1</sup> | <0.001<sup>5</sup> |
| HAVO/VWO (Pre-University Education) | 5 (8.2) |  |  |
| HBO/University | 4 (6.6) |  |  |
| Maternal occupation, n=79 |  |  |  |
| Not involved in IGA | 40 (50.6) | 48.0<sup>2</sup> | 0.82 |
| Involved in IGA | 39 (49.4) | 52.0<sup>2</sup> |  |
| Paternal occupation, n=75 |  |  |  |
| Not involved in IGA | 4 (5.3) | 17.0<sup>2</sup> | <0.001<sup>3</sup> |
| Involved in IGA | 71 (94.6) | 83.0<sup>2</sup> |  |
| Monthly family income, n=78 |  |  |  |
| <1500 SRD (<150US) | 19 (24.3) | N/A | N/A |
| 1500-3000 SRD (150-300 USD) | 33 (42.3) | N/A |  |
| >3000 SRD (>300 US) | 26 (33.3) | N/A |  |
| Household with electricity, n=80 |  |  |  |
| Yes | 78 (97.5) | 97.4<sup>1</sup> | 0.47<sup>3</sup> |
| No | 2 (2.5) | 2.6<sup>1</sup> |  |
| Household with water supply, n=80 |  |  |  |
| Yes | 74 (92.5) | 97.5<sup>1</sup> | 1.0<sup>3</sup> |
| No | 6 (7.5) | 2.5<sup>1</sup> |  |
| Number of persons per room used for sleeping, mean (SD), 95% CI | 2.1 (1.1), 1.9-2.4 | 1.9<sup>2</sup> | <0.001<sup>6</sup> |

<sup>1</sup> MICS 2018 (17); <sup>2</sup> ABS Statistics Suriname 2018 Households (42); <sup>3</sup> Fisher's exact test; <sup>4</sup> Chi-squared test; <sup>5</sup> binomial test; <sup>6</sup> One-sample t test; IGA Income generating activities

### Clinical description and motor function of participating children

**Table 2** summarizes the clinical description and motor function of participating children with CP in SUR-CPR. Among all children, 73.2% had spastic CP, 7.3% had dyskinesia, 12.2% had mixed type of CP and type of CP remained unknown for 7.3%. Among children with spastic CP, the majority (63.6%) had bilateral CP. Of all children, 58.8% had GMFCS level III-V, 40.6% had MACS level III-V, 48.7% had CFCS level III-V and 32.9% had EDACS level IIIV. At least one associated impairment was present in 46.3% children. The median age at diagnosis of CP was 5y 2mo, and 59.7% had a first confirmed diagnosis of CP at the time of registration.

**Table 2:** Clinical description and motor function of participating children.

| Clinical characteristics | Total, n (%) |
| --- | --- |
| **Predominant motor type, n=82** |  |
| Spasticity | 60 (73.2) |
| Dyskinesia | 6 (7.3) |
| Mixed type | 10 (12.2) |
| Unknown | 6 (7.3) |
| **Topography, n=55** |  |
| Unilateral | 18 (32.7) |
| Bilateral | 35 (63.6) |
| Unknown | 2 (3.6) |
| **GMFCS Level, n=80** |  |
| I | 22 (27.5) |
| II | 11 (13.8) |
| III | 5 (6.3) |

| **Clinical characteristics** | **Total, n (%)** |
| --- | --- |
| IV | 6 (7.5) |
| V | 36 (45.0) |
| **MACS level, n=74** |  |
| I | 29 (39.2) |
| II | 15 (20.3) |
| III | 2 (2.7) |
| IV | 7 (9.5) |
| V | 21 (28.4) |
| **CFCS level, n=74** |  |
| I | 27 (36.5) |
| II | 11 (14.9) |
| III | 11 (14.9) |
| IV | 9 (12.2) |
| V | 16 (21.6) |
| **EDACS level, n=76** |  |
| I | 44 (57.9) |
| II | 7 (9.2) |
| III | 11 (14.5) |
| IV | 13 (17.1) |
| V | 1 (1.3) |
| **Presence of at least one associated impairment, n=82** |  |
| No | 44 (53.7) |
| Yes | 38 (46.3) |

### Timing and probable causes of brain injury/ CP among participating children

Among all children, 82.5% had pre- or perinatal causes of CP and the other 17.5% had postneonatal causes. Of the CP cases acquired post-neonatally, the majority had infectious causes of origin. **(Table 3)**

**Table 3:**
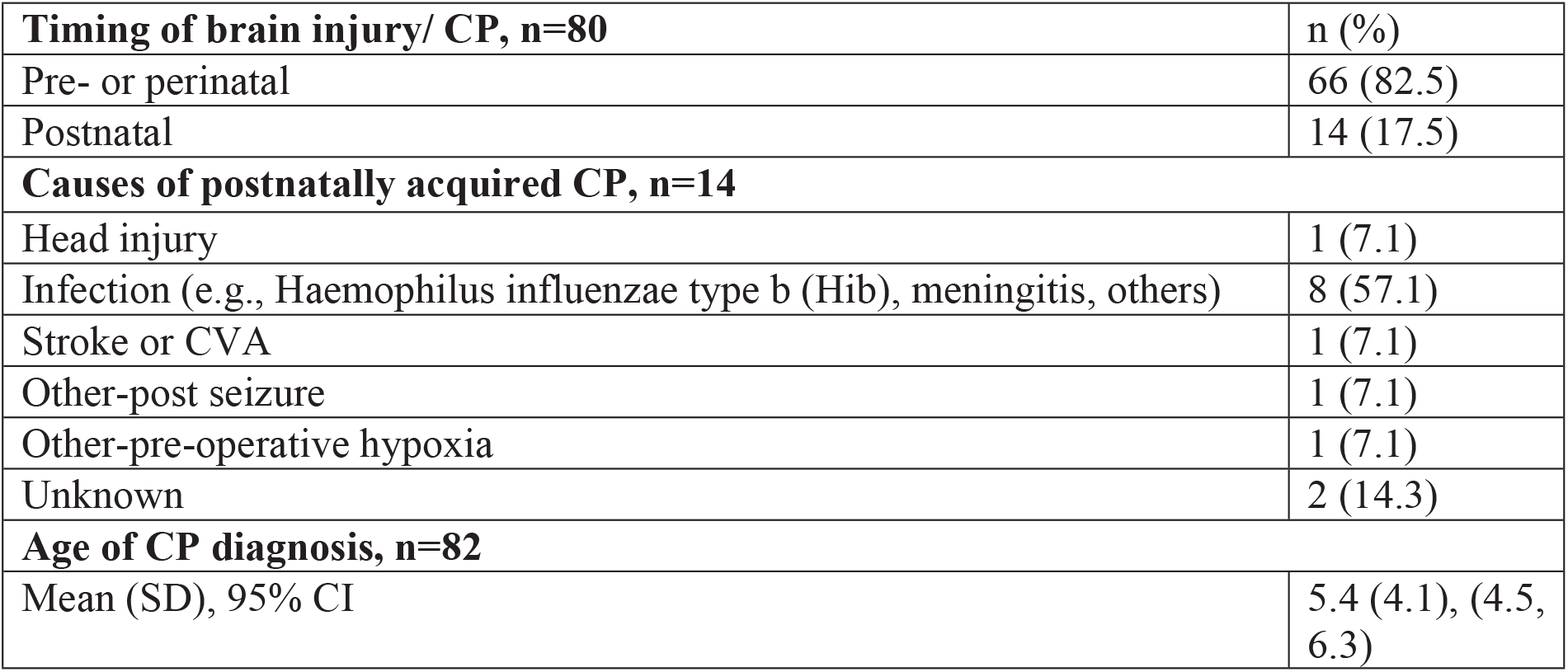
Timing of brain injury and causes of CP.

### Known risk factors

The proportions of different known risk factors among children with CP are summarized in **Table 4**. Most mothers (92.4%) had received antenatal check-ups (ANC) during their pregnancy with the children with CP. Among all children, 42.2% had preterm birth (mean (SD) gestational age: 35.9 (4.1) weeks) compared to the national average (13.9%) (*p*<0.001). Birthrelated complications were reported significantly higher (57%) than the national average (15.0%) (*p*<0.001). Furthermore, 50% of those who reported birth-related complications had preterm birth. Low-birth weight (LBW) was significantly higher among children with CP (40.0%) than reported nationally (15.1%) (*p*=0.01). Thirty nine percent of children had signs of birth asphyxia. Nearly two-third (61.7%) of the children in our cohort were admitted to Neonatal Intensive Care Unit (NICU), and of them, 54.0% required ventilation support.

**Table 4:**
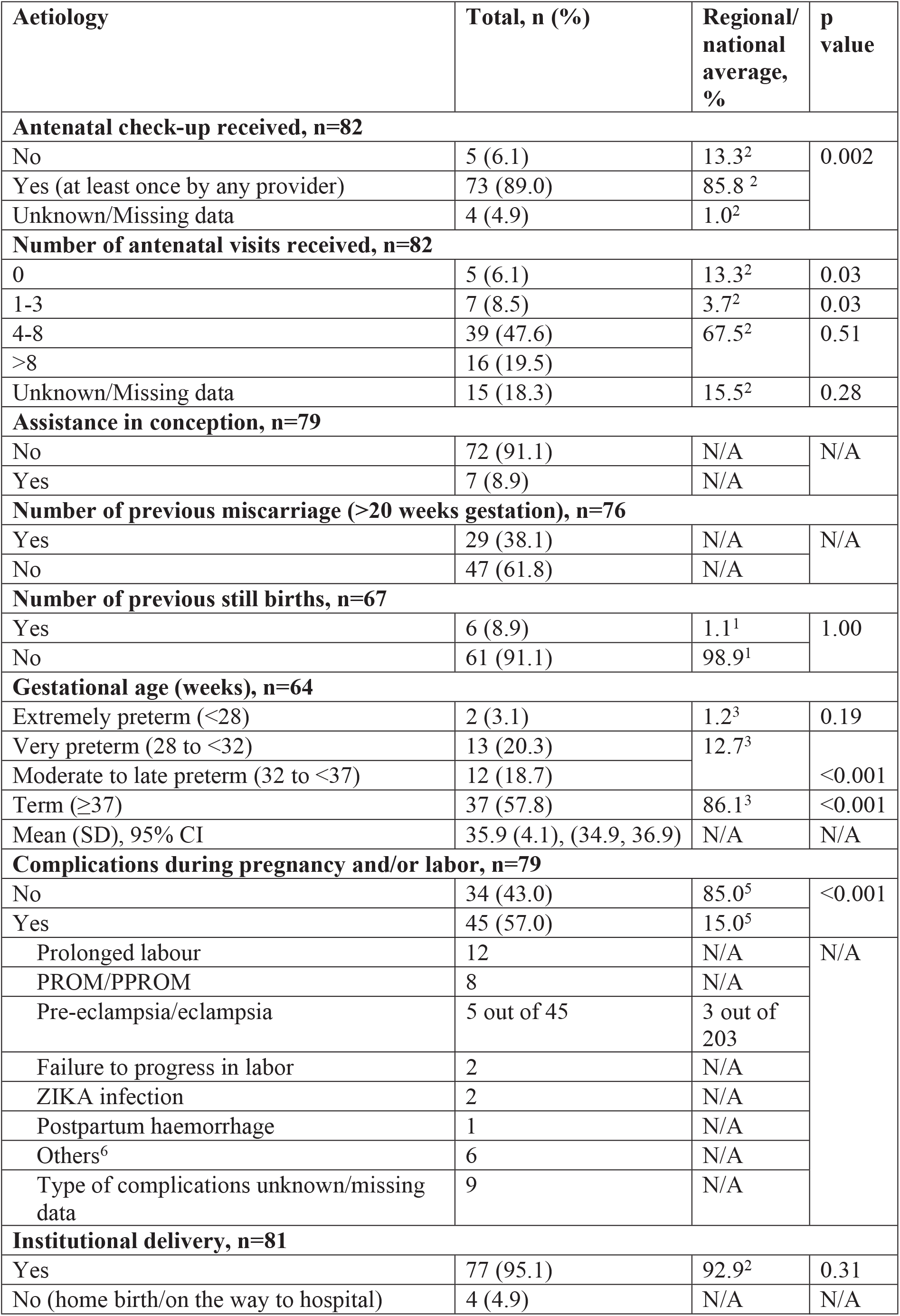
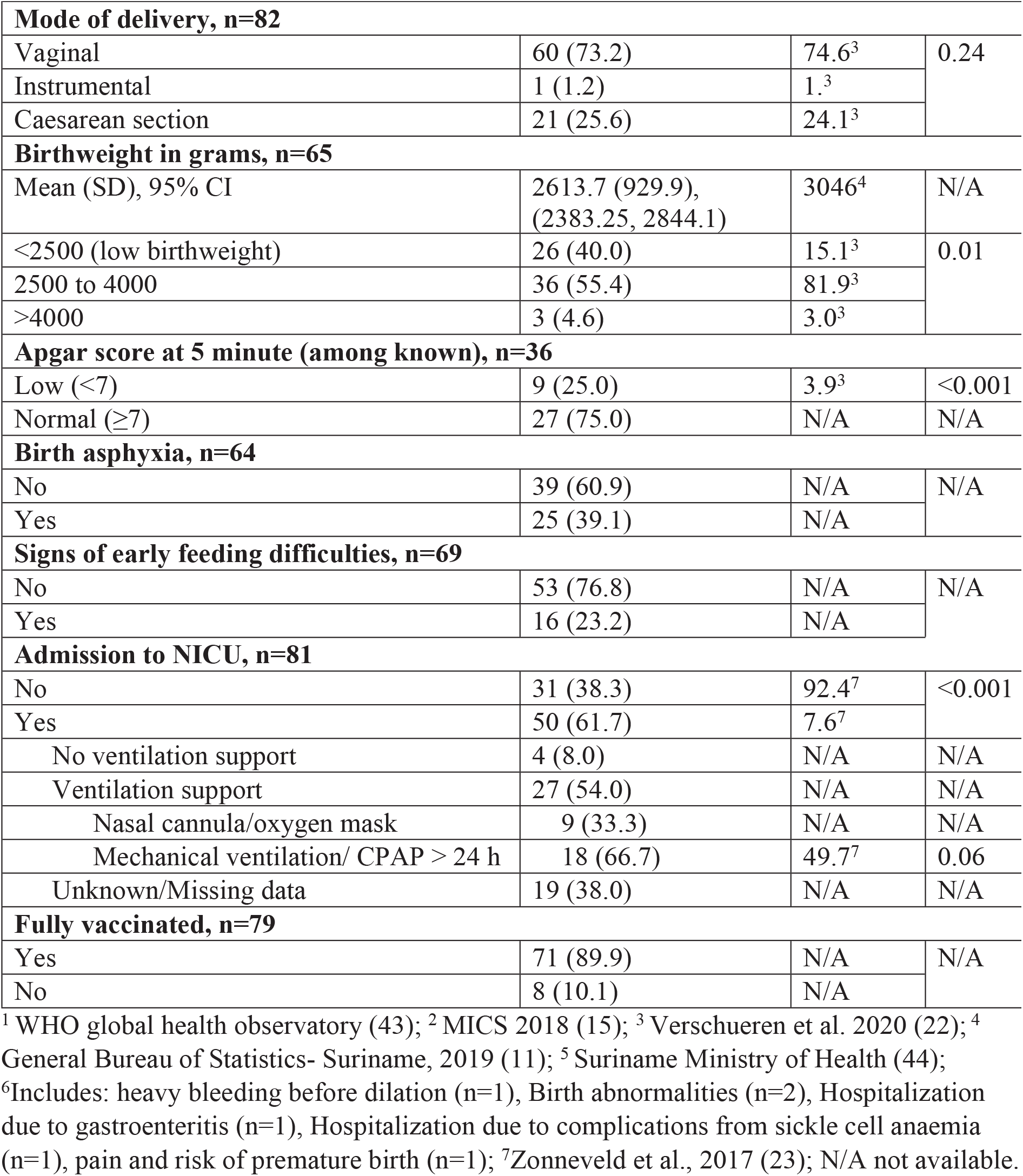
Aetiology/ risk factors of CP among participating children.

### Rehabilitation and education status

The rehabilitation and educational status of participating children is summarized in **Table 5**. Among all children, 15.9% had never received any rehabilitation services. Among the others (i.e. who received rehabilitation at least once), physical therapy was the most commonly received service (84.1%), followed by speech and language therapy (20.6%). However, of those who ever received rehabilitation services, more than half (56.7%) had not received any services in the past six months prior to registration into SUR-CPR. Only 28.5% of the children with GMFCS level IV-V had received a wheelchair.

**Table 5:** Rehabilitation status of participating children.

| **Rehabilitation status** | **n (%)** |
| --- | --- |
| **Ever received any therapy, n=76<sup>1</sup>** |  |
| No | 13 (17.1) |
| Yes | 63 (82.9) |
| **Type of therapy (ever received)<sup>2</sup>** |  |
| PT | 53 (84.1) |
| OT | 7 (11.1) |
| SLT | 13 (20.6) |
| **Received any therapy in 6 months prior registration assessment, n=60<sup>2,3</sup>** |  |
| No | 34 (56.7) |
| Yes | 26 (43.3) |
| **Type of services received in past 6 months<sup>2</sup>** |  |
| PT | 24 (92.3) |
| OT | 4 (15.4) |
| SLT | 8 (30.7) |
| **Ever received any assistive devices, n=77** |  |
| No | 53 (68.8) |
| Yes | 24 (31.2) |
| **Type of assistive devices received, n=24** |  |
| Wheelchair | 15 (62.5) |
| Walking aid | 3 (12.5) |
| Stroller | 1(4.2) |
| Standing frame and walking aid | 1 (4.2) |
| Custom made footwear | 2 (8.3) |
| Hearing aid | 1 (4.2) |
| Custom made seat and stroller | 1 (4.2) |
| **School attendance and type (among children aged ≥4y), n=47** |  |
| None | 15 (31.9) |
| Special education/schools | 14 (29.7) |
| Mainstream schools | 11 (23.4) |
| Day care | 7 (14.9) |

<sup>1</sup> Missing data; <sup>2</sup>Not mutually exclusive; <sup>3</sup> Among children who ever received services

Among school-aged children (i.e. aged 4 years or above), 31.9% were not enrolled in any school, 23.4% were enrolled in mainstream schools and 29.7% were enrolled in special education programs/schools. The common reasons (not mutually exclusive) for not attending school were parental refusal due to the child being different (n=4), school distance and transport-related problems (n=3), lack of inclusive education available in the locality (n=2) and financial problems (n=2).

## Discussion

In this study, we described the epidemiology of CP among children in Suriname using data from the Suriname CP register i.e., SUR-CPR. To the best of our knowledge, this is the first CP register established in South America. The SUR-CPR was established with the aim of setting up a platform for the national CP register using the existing health system infrastructure in the country. So that data from the CP register could generate important evidence on the epidemiology of CP (e.g., clinical description, aetiology, risk factors) and facilitate translational research to improve functional and health related outcome of children with CP in Suriname. The preliminary findings reported here are therefore crucial and have strong implications for future research and provide evidence-based intervention and services for children with CP in 0the country. Although we limited our reporting to descriptive analysis only, due to the small number of participants, the detailed methods and data reported will form the basis for integrating and scaling-up of the CP register activities into the existing health system. Findings from this will also facilitate future studies on potential interventions for the prevention or reduction of the burden of known risk factors of CP in Suriname. As observed in another LMIC (i.e., Bangladesh) (20), continuation of the set-up and registry (i.e., SUR-CPR), might also raise awareness and help to implement the guidelines for early diagnosis and consequently early intervention for children with CP in Suriname.

Unlike other population-based surveillance of children with CP from LMICs, the majority of the children registered into the SUR-CPR are aged less than 10 years. This difference could be attributed to potential recruitment bias (hospital vs. population-based) as well as the low survival probability of older children with CP, which have been reported in other CP registers from LMICs. (31, 32) The predominance of young children found in our study is consistent with other institution-based LMIC CP registers e.g., Vietnam and Ghana. (6, 7) In addition to the demographic differences, we observed a significantly lower educational attainment among parents of children with CP in Suriname when compared with the national average. (11) A similar pattern was observed in other LMICs. (4-7) Low maternal education has been previously identified as a predictor of malnutrition, and non-receipt of rehabilitation services among children with CP in LMICs as well as among the general population in Suriname. (4, 7, 32) Unfortunately, this paper could not explore the relationship between these characteristics due to the low number of case ascertainment. Furthermore, we observed that the monthly household income of the registered children was substantially lower than the national average (∼410 USD per capita in 2020) in Suriname. The relationship between income (and/or poverty) and pre-/perinatal care, health, nutrition, rehabilitation and education of children with CP specially in LMICs has been documented in several studies in the past. (4, 6, 7, 32, 33) However, this difference in income should be interpreted with caution as the national data mostly represents the gross salary of civil servants.

Nevertheless, as described previously, this indicates that children with CP from such financially disadvantaged families may struggle to access diagnosis and rehabilitation services as the govt.subsidized health care (i.e., health insurance) partially supports such specialist services. Moreover, while accessing GP and medical specialists is covered by the state health insurance package, accessing services from the PT, OT and ST are only partly covered in this package (e.g., 60% of 10 physical therapy sessions is refunded yearly per person). This complex system is likely to increase the risk of lost to follow-up and non-uptake of the referral, thus delaying the scope for early diagnosis and early intervention for children with CP and other neurodevelopmental disorders in Suriname. The percentages of commonly known risk factors of CP such as preterm birth and low birth weight were also comparatively higher among children registered in the SUR-CPR than reported nationally in Suriname. (22) A similar rate of preterm birth (43%) and low birth weight (43%) was reported among children with CP in Australia. (8) The burden was also higher when compared to findings from a hospital-based surveillance of children with CP in Vietnam. (7) A recent study that reported a similar high burden of preterm birth and low birth weight among women living in the tropical rain forest of Suriname showed a significant association between the ethnicity of the mothers, their ANC visits and birth outcome. (34) A large number of mothers in our cohort reported pregnancy and/or birth-related complications (including obstructed/prolonged labor, pre-eclampsia/ eclampsia, PPH). Furthermore, signs of birth asphyxia and low APGAR score were also highly prevalent among the participating children with CP. A similar high burden was reported in other LMICs. (6, 7) In absence of essential emergency obstetric and newborn care (EmONC), these complications increase the risk of morbidity/injury and mortality of both mothers and the fetus/ newborn. Our data also show that a high proportion of children with CP in our cohort required NICU support, and this percentage was higher when compared with another recent study in Suriname and data from a hospital-based surveillance program of CP in Vietnam. (7, 23) This could be due to potential recruitment bias. Although in Suriname the majority of births take place at institutions, there are only six tertiary hospitals that support advanced health care needs (including NICU support) of the entire population in the country, and of those five, four are located in the capital (including the tertiary hospital where SUR-CPR has been established). It is likely that pregnant mothers and/or newborns in the rural part of the country have comparatively less access to those EmONC services and are likely to be referred to the tertiary hospitals in the capital. Our data also show that a high proportion of children with CP required NICU support, and this percentage was higher when compared with another recent study in Suriname and data from a hospital-based surveillance program of CP in Vietnam. (7, 23) Evidently these children are considered as high-risk infants with poor neurodevelopmental outcome. More intensive and close follow-ups including the trial of novel therapeutics for these high-risk NICU graduates in LMICs like Suriname could create opportunities for early diagnosis of CP and enable access to early interventions. In our cohort, the majority of children had pre- or perinatally acquired CP, similar to findings from other LMICs and HICs. (4, 6, 7, 35) However, the mean age at diagnosis was delayed when compared with other institution-based CP registers in LMICs (e.g., Vietnam and Ghana) (4, 6, 7) The delayed diagnosis of CP could be attributed to the lack of early detection services. at the community level and non-uptake of the referral to tertiary health facilities for specialized assessments as discussed previously. This consequently delays the opportunity for early intervention of those children with CP in Suriname. Given the high proportion of institutional birth in Suriname, a screening system following international guidelines could be established to identify children at risk of developing CP. Similarly, screening services to identify high-risk infants could be integrated at NICU and subsequent follow-ups for further assessments e.g., general movement assessment and using standard tools could be scheduled/ adopted to promote early detection of CP among children in Suriname. (36) Similar to other LMICs, (6) the majority of postnatally acquired CP in Suriname were due to infectious causes. Preventive approaches including compliance with immunization during pregnancy and early childhood should be prioritized. The majority in our cohort had spastic bilateral CP and/or severe motor function limitations (i.e. GMFCS and MACS level III-V). The findings are consistent with data reported from institution-based settings in Vietnam and Ghana, however, are contrary to HICs such as Australia, Sweden, and Denmark. (4, 6, 7, 37, 38) A high burden of eating and drinking difficulties has been found in our study. The overrepresentation of EDACS level III-V is likely to be associated with the high prevalence of GMFCS level III-V. (39) Delayed CP diagnosis and lack of early intervention and rehabilitation services is likely to increase the severity of functional impairment among those children.

Nevertheless, more than half of the children did not receive any rehabilitation in the past six months preceding the registration even though those children were recruited from the paediatric unit of a tertiary hospital (i.e. AZP). Furthermore, the majority received PT only, and nearly two-thirds of children with GMFCS IV-V did not receive any assistive device to help them with mobility. A recent study from Bangladesh reported that 96.3% of children with CP and GMFCS level III-V have never received assistive devices. (40) Almost 20% of the children across different GMFCS levels had never attended any therapy; this is concerning specially because the data were collected from tertiary health facility where children are referred for detailed screening and rehabilitation services, indicating that the situation may be worse in communitybased settings. Such poor rehabilitation status could be attributed to multiple factors including care pathways not addressing the lack of knowledge, transportation difficulties or financial support (since insurance companies only provide 10 treatment sessions yearly). The low proportion of children enrolled in mainstream schools is also consistent with findings from other LMICs. (6) In a recent study, the foundational reading and numeracy skills were also reported to be low among children with functional disabilities when compared to children without disabilities in Suriname. (41) Furthermore, findings from the SUR-CPR suggest that parental refusal due to the child being different, school distance, transport-related and financial issues are key underlying causes of non-attendance to schools among registered children. The findings are consisted with those reported in a few other LMICs (e.g., Bangladesh, Vietnam, Nepal). Advocacy and awareness raising programs highlighting the need of inclusion of children in daily activities including inclusive education could improve the school attendance of children with CP in Suriname. Training of more special educators, allocating more resource to upgrade the mainstream schools with necessary supplies and materials would facilitate inclusiveness, as well enable children with CP to get the education and training required for future prospects and a better-quality life in LMICs like Suriname.

Despite considerable effort, the current study has several limitations including a chance of recruitment bias due to the nature of the surveillance setting, thus the findings should be interpreted with caution. However, in Suriname, data on childhood disabilities or functioning status are collected in four to ten years intervals (e.g., the general Bureau of Statistics holds a Census throughout the country every eight to ten years, the latest census was carried out in 2012; whereas MICS is usually conducted in every four years). Both lack information on CP diagnosis or the epidemiology. Findings from this research therefore provides important data for understanding of the status, future research and evidence-based services for children with CP in the country. Although we strictly followed a standard guideline and case definition for diagnosis and registration of children with CP in the SUR-CPR, we had different assessors for documentation of the motor classifications, which may have posed mild risk with low inter-rater reliability in a few children registered into the SUR-CPR. Finally, part of the data was collected based on parent reporting which could be affected by recall bias and missing data.

## Conclusion

As mentioned earlier, to our knowledge the SUR-CPR is first of its kind in Suriname and the South American countries, thus reporting one of the first detailed epidemiological profile of CP in children from the region. Although current data is not generalizable to all children with CP in Suriname, through this research we have established the methods/platform for a database of children with CP which can be scaled-up to community-settings by partnering and strengthening the existing health system referral networks (e.g., 50 health centres of Medical Mission, 50 clinics of RGDs) in Suriname. The findings contribute crucial data to the current knowledge base specially to understand the CP epidemiology in different LMICs and subsequently the global picture of CP. In Suriname, a large proportion of children with CP had known risk factors with comparatively delayed age of diagnosis and severe motor impairments. Furthermore, one-third of the children were not enrolled in any education system and the majority did not receive comprehensive rehabilitation services for better functional outcomes. The preliminary findings from this research have portrayed the CP epidemiology and could be used to advocate for the high service needs, facilitate early diagnosis, and improve access to early intervention, rehabilitation and education services for children with CP in Suriname. It is anticipated that the ongoing engagement and activities of the surveillance system will create awareness among health care and rehabilitation professionals, highlight the needs for integrating and expanding the SURCPR services to the public health facilities of the RGD and MM, which will eventually promote early diagnosis and early intervention following international guidelines for children with CP in Suriname.

## Data Availability

All data produced in the present study are available upon reasonable request to the authors

## Funding and Acknowledgements

Training of clinicians and the set-up of the SUR-CPR was supported by Cerebral Palsy Alliance

Research Foundation Project Grants (PRG02919 & PRG10318).

## Notes

### Competing Interest Statement

The authors have declared no competing interest.

### Author Declarations

Medical Ethical Committee of the Ministry of Health, Suriname (VG34-17A)

### Summary of Updates

Error in the author list

